# Cortical activation and motor-cognitive performance during dual-task walking across healthy aging and Parkinson’s disease: a standardized longitudinal fNIRS and gait analysis protocol

**DOI:** 10.64898/2026.06.02.26354715

**Authors:** Luiza de Mattos Aranha, Paloma Rodrigues da Silva, Denilson Feijoeiro Garcia, Larissa Bitarães Rodrigues dos Santos, João Ricardo Sato, Gabriel Venas Santos, Kelly Rosa Braghetto, Maria Elisa Pimentel Piemonte.

**Affiliations:** Neuroscience and behavior department, Psychology Institute, University of São Paulo, São Paulo, Brazil; Department of Physiotherapy, Speech Therapy and Occupational Therapy, Faculty of Medicine of University of São Paulo, São Paulo, Brazil; NeuroMat Teaching, Research and Dissemination Center, Institute of Mathematics and Statistics, University of São Paulo, São Paulo, Brazil; Center of Mathematics, Computing, and Cognition, Federal University of ABC, São Bernardo do Campo, São Paulo, Brazil

**Keywords:** Aging, Dual-task, Gait, Automaticity, Parkinson’s disease, fNIRS

## Abstract

**BACKGROUND:** Aging and Parkinson’s disease (PD) reduce gait automaticity and increase cognitive demand during walking. Although dual-task (DT) paradigms investigate cognitive–motor interference, evidence remains limited by heterogeneous tasks, predominant focus on prefrontal cortex (PFC) activity, and variability in functional near-infrared spectroscopy (fNIRS) methods. This study investigates whether longitudinal changes in cortical activation during DT walking differ among young adults, older adults, and individuals with PD, and how these changes relate to DT costs over 5 years.

**METHODS:** This longitudinal observational study follows STROBE and fNIRS guidelines and will be conducted in a controlled laboratory (Rede Amparo, CEPID NeuroMat, University of São Paulo). Participants will be evaluated annually under three randomized conditions: motor single-task walking, cognitive single-task phonemic verbal fluency and DT walking with phonemic verbal fluency, each repeated 10 times. The primary outcome measure will be longitudinal changes in cortical activation during DT walking, quantified by oxygenated hemoglobin (HbO) signals measured with fNIRS in prefrontal and premotor cortical regions. The main predictors of interest will be motor and cognitive DT costs. Covariates will include age, sex, education, cognition, balance, mood, and disease severity in the PD group. Spatiotemporal gait parameters, including gait speed, step length, stride length, step time, base of support, double support, stance phase, and variability, will be recorded using the GAITRite® system, and DT costs will be calculated for selected parameters. Cortical activation will be assessed using a 66-channel wearable fNIRS system with short-separation channels.

**DISCUSSION:** By combining randomized task blocks, separate motor and cognitive conditions, broader cortical coverage, and concurrent neural and gait assessment across three groups annually, this protocol is expected to provide a comprehensive characterization of cognitive–motor interference during walking and its evolution, supporting interpretation of cortical and behavioral responses. The study may help distinguish age-related adaptations from PD-specific alterations and clarify whether increased cortical recruitment during DT gait reflects compensation, reduced neural efficiency, or ceiling effects, refining understanding of gait automaticity decline and informing rehabilitation and non-invasive brain stimulation approaches.

**Administrative information:** The numbers in parentheses in this protocol refer to STROBE checklist item numbers. The order of the items has been modified to group similar items.

## INTRODUCTION

### BACKGROUND

Aging, through its combined motor and cognitive effects, contributes to a progressive reduction in gait automaticity and to greater cognitive-motor interference during walking [1,2,3]. Age-related changes in gait and mobility include reduced adaptability of locomotor control and increased vulnerability to divided-attention demands [1,4], while cognitive changes affecting processing speed, attention, and executive function further increase reliance on cortical control during locomotion [2,3]. In Parkinson’s disease (PD), these alterations are further amplified by neurodegenerative processes affecting basal ganglia-cortical circuits, resulting in greater disruption of automatic gait control, increased attentional demands during walking, and more pronounced impairments under dual-task conditions [5,6,7].

Walking in everyday life rarely occurs in isolation. In most real-world situations, gait is performed concurrently with other motor or cognitive activities, such as talking, planning, monitoring the environment, carrying objects, or responding to unexpected stimuli. For this reason, dual-task and multiple-task walking paradigms provide an ecologically relevant model for investigating locomotor control under conditions that more closely resemble daily life [4,8,9]. In both healthy aging and PD, walking under divided-attention conditions is associated with greater gait deterioration than single-task walking, including reductions in speed and step length, increased variability, and poorer postural stability [5,10,11]. These changes are clinically relevant because they are associated with greater functional vulnerability and increased risk of falls, particularly in healthy older adults and in people with PD, for whom safe locomotion often depends on the ability to manage competing cognitive and environmental demands [7,8].

In response to these increased task demands, cortical activation during walking is modified, reflecting greater reliance on supraspinal control mechanisms when gait automaticity is reduced [3,12,13]. Previous fNIRS studies have shown task-related increases in prefrontal activation during dual-task walking in young and older adults, while studies in PD suggest even greater dependence on cortical control during complex walking conditions [13,14,15]. However, the functional meaning of this increased activation remains insufficiently understood. Greater cortical recruitment may reflect adaptive compensation, reduced neural efficiency, task-related ceiling effects, or distinct neural strategies across populations, depending on the specific task and the characteristics of the individuals being studied [3,5,16]. As a result, it remains unclear how changes in cortical activation during dual-task walking relate to concurrent motor and cognitive performance, or whether greater recruitment of these regions helps attenuate the negative effects of dual-tasking on gait and cognition.

This uncertainty is likely due, at least in part, to important methodological limitations in existing literature. Previous studies have used highly heterogeneous cognitive tasks, engaging different domains and levels of executive demand, which limits comparability across findings [9,17,18]. Many investigations have relied on pairwise group comparisons rather than examining young adults, older adults, and individuals with PD within a single standardized framework [5,10,13]. In addition, most studies have focused predominantly on prefrontal activation, providing only a partial view of the cortical networks involved in gait control [12,15,16]. Other recurring limitations include variability in fNIRS acquisition and processing procedures, insufficient standardization of experimental design, and limited ability to directly relate neural measures to concurrent motor and cognitive performance in a consistent manner across studies [15,19,20]. An additional limitation is the scarcity of longitudinal studies capable of showing how aging and PD progression influence changes in cortical activation during dual-task walking over time. Available longitudinal evidence has mainly focused on behavioral dual-task gait outcomes in older adults, including prediction of falls and cognitive decline [21,22,23] or on longitudinal changes in brain function during dual-task locomotion in healthy older adults assessed with fNIRS [24,25]. By contrast, longitudinal studies integrating repeated cortical measures during dual-task walking in PD or directly comparing trajectories of healthy aging and PD under the same experimental framework, remain limited. Together, these limitations restrict the interpretation of how cortical recruitment during dual-task walking relates to the magnitude of cognitive–motor interference and to the preservation or deterioration of motor and cognitive performance.

To address these gaps, the present protocol adopts a standardized and methodologically rigorous framework designed to support more reliable investigation of cognitive–motor interference across different populations. By applying the same experimental paradigm to young adults, healthy older adults, and individuals with PD, the protocol enables direct comparison of cortical and behavioral responses under equivalent task demands, reducing the interpretive limitations of pairwise or non-comparable study designs [5,10,13]. The inclusion of separate motor and cognitive single-task conditions provides a more robust baseline for interpreting dual-task-related changes in both cortical activation and performance, allowing clearer distinction between neural activity related to gait, cognition, and their interactions [13,14]. In addition, the use of 10 repeated blocks per condition increases the stability of both behavioral and hemodynamic estimates, which is particularly relevant in fNIRS studies given the susceptibility of the signal to motion artifacts and trial-to-trial variability [20,26]. The choice of phonemic verbal fluency as the secondary task also represents a methodological advantage, as it imposes meaningful executive demand while remaining feasible across a broad functional spectrum, including young adults, healthy older adults, and individuals with PD [17,18,27]. To further reduce potential learning and order effects, task blocks will be presented in randomized order across trials. Finally, the protocol combines broader cortical coverage, including prefrontal and premotor regions, with simultaneous assessment of multiple spatiotemporal gait parameters and relevant clinical variables, thereby providing a more comprehensive framework for examining how patterns of cortical recruitment relate to motor and cognitive performance during dual-task walking [12,19,28].

Taken together, this protocol is designed to address a central unresolved question in the field: how changes in cortical activation during dual-task walking relate to concurrent motor and cognitive performance across healthy aging and PD. By reducing key methodological limitations of previous studies and integrating neural, motor, and cognitive measures under standardized conditions, this approach may advance understanding of the mechanisms underlying reduced gait automaticity and provide a framework that can also support future longitudinal investigations of age- and disease-related trajectories. In doing so, it may help identify clinically meaningful neural signatures and inform future rehabilitation strategies and non-invasive brain stimulation approaches grounded in these mechanisms.

### OBJECTIVES

To investigate longitudinal changes in cortical activation during dual-task walking across young adults, healthy older adults, and individuals with PD, and to examine how these changes relate to motor and cognitive dual-task costs, over 5 years.

## METHODS

### STUDY DESIGN

This is a longitudinal study that follows all the guidelines of the Strengthening the Reporting of Observational Studies in Epidemiology (STROBE) Statement and recommendations of consensus guide to using functional near-infrared spectroscopy (fNIRS) in posture and gait research [20].

### ETHICS STATEMENT

The protocol was developed following the Strengthening the Reporting of Observational Studies in Epidemiology (STROBE) and was approved by the Research Ethics Committee of the Hospital das Clínicas, Faculty of Medicine, University of São Paulo (CAAE: 67388816.2.0000.0065; Approval number 6.913.344).

### SETTING

The study will be conducted in the research laboratory of REDE AMPARO, part of CEPID NEUROMAT (www.neuromat.numec.prp.usp.br). The laboratory is accessible, flat, isolated from excessive noise, and has controlled temperature and lighting.

### PARTICIPANTS

A convenience sample will be recruited through REDE AMPARO (http://www.amparo.numec.prp.usp.br). Eligibility criteria for healthy young and older adults will include: the absence of any diagnosed neurological disorder and walking independently. (i.e., without an assistive device or another person’s assistance). Eligibility criteria for individuals with PD will include a diagnosis of idiopathic PD confirmed by a neurologist, use of dopaminergic medication and walking independently (i.e., without an assistive device or another person’s assistance). Exclusion criteria for all groups will include a diagnosis of dementia, musculoskeletal or cardiorespiratory disorders, as well as uncorrected visual or auditory impairments that could interfere with test performance.

### VARIABLES

Participants will undergo annual in-person assessments over 5 years, conducted by specialized therapists. Demographic data (date of birth, age, gender, education and profession) will be collected during the first assessment.

#### Primary Outcomes Cortical Activity

The primary outcome measure will be longitudinal cortical activation during dual-task walking across prespecified prefrontal and premotor cortical regions. Cortical activation will be indirectly assessed using fNIRS, a noninvasive neuroimaging technique that measures changes in oxygenated (HbO) and deoxygenated hemoglobin (HbR) concentrations, providing an indirect index of neuronal activity [31]. A wearable, portable, wireless NIRSport2 continuous-wave fNIRS system (NIRx Medizintechnik GmbH, Berlin, Germany) will be employed, using dual-wavelength illumination (760 nm and 850 nm) to quantify temporal variations in HbO and HbR. HbO will be used as the primary cortical measure because of its greater sensitivity and signal-to-noise ratio in walking paradigms [15,19].

A custom 66-channel probe array - comprising 16 light sources and 30 light detectors with source-detector separations of ∼30 mm - will be mounted on an adjustable head cap according to the international 10–20 EEG system. Placement will be centered at Cz, ensuring standardized alignment across participants, with coverage focused on bilateral PFC and premotor cortices (Fp1, Fp2, F3, F4, and adjacent regions), as shown in Figure 1. Sixteen additional short-separation channels (approximately 15 millimeters distance between sources and detectors) will be included to capture superficial hemodynamics for subsequent regression of extracerebral signals [32]. Prior to the initiation of data acquisition, the fNIRS hardware will be calibrated while the participant maintains an orthostatic posture. A black shower cap will be placed over the head cap to minimize exposure of the optodes to ambient light, thereby reducing potential interference with optical signal acquisition.

**Figure 1.**
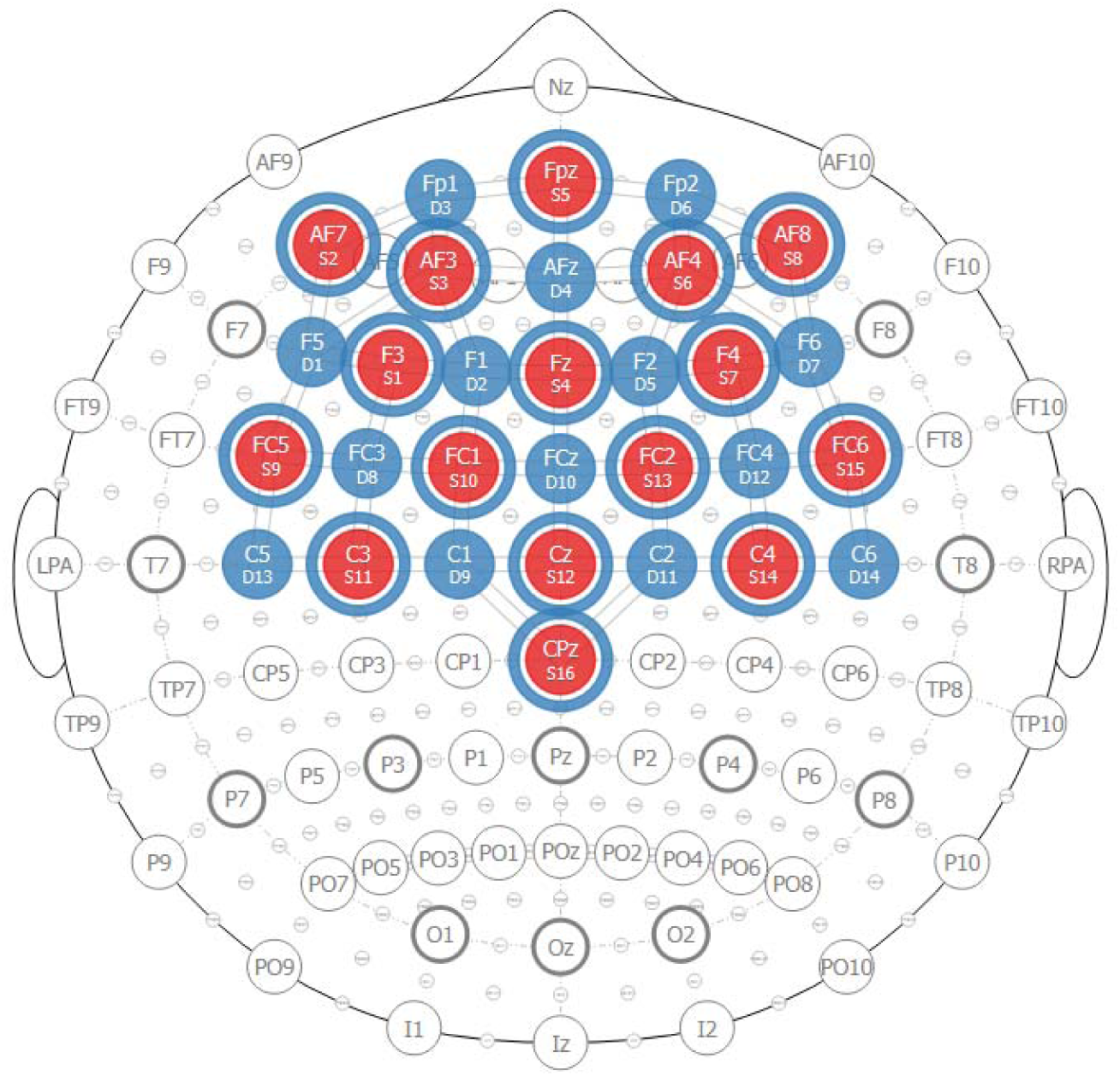
fNIRS optode configuration and positioning. Figure 1 represents the schematic organization of the functional near-infrared spectroscopy (fNIRS) montage showing the spatial distribution of sources and detectors over the prefrontal cortex and premotor regions. Red circles: Sources; Red circles circled in blue: Sources with short-distances detectors; Blue circles: regular detectors.

The head cap optodes will be connected to a portable receiver worn in a backpack, allowing assessments during walking. Data will be sampled at 5.1 Hz and streamed to a laptop running Aurora Software v.2021.4 (NIRx Medical Technologies LLC, Glen Head, NY, USA).

Raw optical signals will be converted to changes in HbO and HbR concentrations using the modified Beer-Lambert law. Preprocessing will include filtering and procedures to reduce physiological and motion-related artifacts (e.g., cardiac and respiratory components), as recommended in prior fNIRS studies [26].

Hemodynamic responses will be analyzed using a general linear model. Task conditions will be modeled as repeated 10-second blocks corresponding to the experimental paradigm, and condition-specific beta coefficients will be estimated for each channel. Group-level analyses will be conducted using individual beta values, accounting for within-subject structure and temporal autocorrelation. For all analyses, HbO signals will be used as the primary cortical measure due to their higher signal-to-noise ratio and greater sensitivity to task-related cortical activation compared to HbR [15,19]. Condition-specific beta coefficients will be estimated at the channel level and then summarized within prespecified prefrontal and premotor regions to generate regional cortical activation measures for longitudinal analyses.

#### Main predictors of interest

The main predictors of interest will be motor and cognitive dual-task costs derived from the experimental protocol. Motor dual-task costs will be calculated from spatiotemporal gait parameters obtained during dual-task and motor single-task walking conditions. Cognitive dual-task costs will be calculated from verbal fluency performance during dual-task walking relative to the cognitive single-task condition. These variables will be used to examine whether longitudinal variations in cortical activation are associated with behavioral performance over time.

#### 1. Gait Performance

The assessment of spatiotemporal gait parameters will be performed using the GAITRite system (GaitRite System, CIR Systems Inc., Franklin, NJ, USA), a portable quantitative gait analysis system that uses a pressure-sensitive walkway to measure spatial and temporal gait parameters [28,33].

Spatiotemporal gait measures obtained during single-task and dual-task walking will be used to derive motor dual-task costs, which will serve as main behavioral predictors in the longitudinal analyses.

Before the beginning of the assessment trials, participants will receive detailed explanations about the procedures, including the conditions to be tested and the required tasks for each condition. After confirming that they understood the instructions and clarifying any questions, a full practice trial will be conducted to promote familiarization and ensure complete comprehension of the task. Data collection will begin only after the participant demonstrates full understanding of all instructions.

It is important to note that, in the DT condition, participants will not receive any instruction regarding which task will need to be prioritized (walking or verbal fluency). The instruction provided will be: “Walk along the walkway at your usual pace while saying as many words as possible beginning with the indicated letter.”

Following the familiarization trial, participants will be positioned at the starting point marked one meter before the beginning of the walkway. After the command, participants will proceed barefoot at their self-selected habitual pace along a straight trajectory, maintaining an upright head posture and visual fixation on an X-shaped marker aligned with eye level on the horizon. They will be then instructed to walk along the GAITRite walkway upon hearing the command “WALK”. When reaching the end of the condition, the command “STOP” will be given, signaling them to interrupt walking. As participants walk along the walkway, the embedded sensors will capture precise spatiotemporal gait parameters, including gait speed, step length, stride length, step time, base of support, double support, and stance phase.

Each participant will perform ten trials for each of the three randomly presented conditions: single-task walking (walking at their usual pace without any concurrent task); cognitive single-task (producing as many words as possible beginning with a designated letter while standing still in an upright position); and DT walking (walking at their usual pace while producing as many words as possible beginning with a designated letter).

The conditions will be presented in a randomized order across trials, with randomization applied within each block to minimize potential order and learning effects.

Each condition will be repeated ten times. Each trial will last 10 seconds and will be followed by a 30-second interval, including rest, repositioning, and preparation phases. The total duration of the experimental protocol will be approximately 20 minutes, as shown in Figure 2.

**Figure 2.**
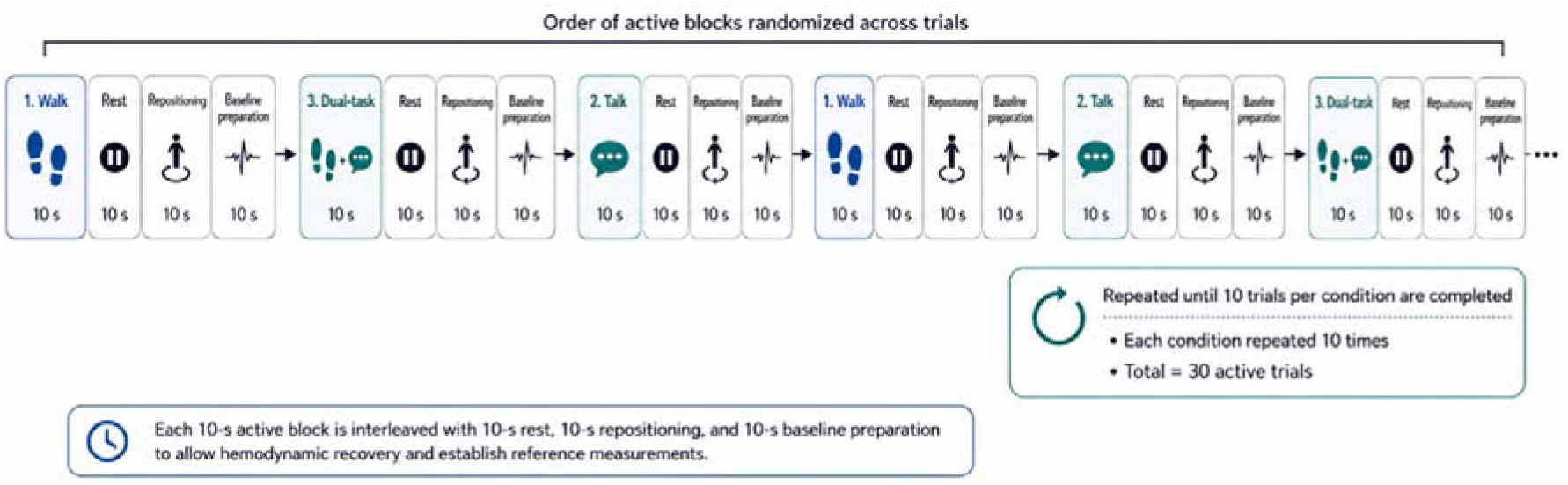
Experimental protocol flowchart. Figure 2 represents the timeline of the experimental conditions, including single-task walking (walk), cognitive single-task (talk), and dual-task walking while talking. Each active block (10 s) will be interleaved with rest periods (10 s), repositioning (10 s) and baseline preparation phase (10 s) to allow hemodynamic recovery and establish reference measurements.

Assessments will be performed simultaneously (fNIRS and GAITRite), ensuring synchronization between motor and neurophysiological variables.

#### 2. Cognitive performance

The verbal fluency task will be used as the cognitive task due to its high cognitive and executive demand and its extensive use in DT paradigms. A verbal fluency task requires continuous lexical retrieval, semantic memory access, and executive monitoring, engaging distributed cortical networks, particularly within left frontal regions, including the dorsolateral prefrontal cortex and inferior frontal gyrus [27]. In addition, tasks with higher executive demand have been shown to induce greater cognitive-motor interference and increased prefrontal activation during walking compared to less demanding paradigms [17,18]. These characteristics make verbal fluency a suitable paradigm to probe the limits of gait automaticity and to examine cortical resource allocation under cognitively demanding conditions.

In standard neuropsychological assessments, verbal fluency tasks are typically administered over 60-second intervals. In the present study, each trial will last 10 seconds and will be repeated 10 times per condition. This adaptation will be implemented to optimize compatibility with the block-design structure required for fNIRS data acquisition, allowing repeated measurement of cortical hemodynamic responses while minimizing fatigue and motion-related artifacts.

#### Cortical Activity and Gait Analysis Evaluation Across Task Conditions

To examine cortical and gait responses under different motor and cognitive demands, cortical hemodynamic activity will be monitored using fNIRS while gait performance is simultaneously assessed using GAITRite (Figure 3), in a block design type, during alternating execution of 10 repetitions of the three experimental conditions. Participants will complete familiarization trials under all experimental conditions prior to the commencement of testing. The total duration of the experimental procedure will be 20 minutes, during which participants will be instructed to report any instances of perceived discomfort.

**Figure 3.**
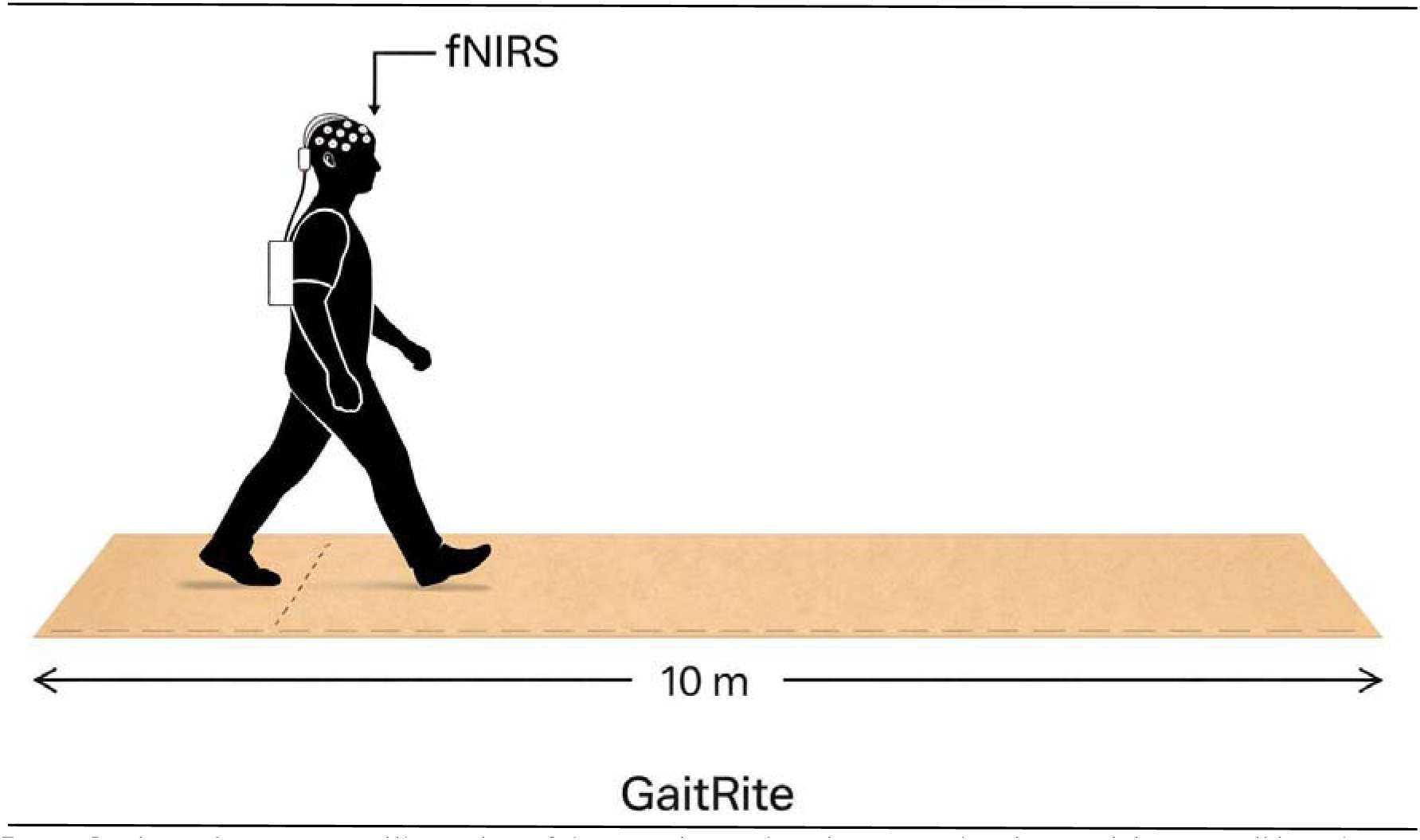
Overground walking setup integrating GAITRite and fNIRS. Figure 3 presents an illustration of the experimental environment showing participants walking along a 10-m GAITRite electronic walkway while wearing a portable functional near-infrared spectroscopy (fNIRS) system.

#### Single-Task Walking

This condition will involve walking at a self-selected habitual speed on the walkway, without any concomitant cognitive demand, for 10 seconds. It is designed to examine cortical activity exclusively associated with control of gait.

#### Cognitive Single-Task

In the cognitive single-task condition, participants will perform a phonemic verbal fluency task while standing. At the beginning of each trial, a distinct target letter will be presented, varying across trials to minimize repetition and learning effects. Participants will be instructed to generate as many words as possible within a 10-second interval.

#### Dual-Task Walking Condition

In the dual-task walking condition, participants will perform the phonemic verbal fluency task while walking at a self-selected speed. At the beginning of each trial, a new target letter will be randomly selected from the same predefined set of letters used in the cognitive single-task condition. The order of letter presentation will be independently randomized for both conditions to minimize potential learning effects and order-related biases. Participants will not be instructed to prioritize either the motor or the cognitive task, allowing for natural allocation of attentional resources between gait and verbal performance.

#### Covariates

The following variables will be included as covariates due to their potential influence on dual-task performance and cortical activation.

#### Montreal Cognitive Assessment [MoCA]

Cognition will be assessed through MoCA, a widely used screening instrument for detecting subtle cognitive deficits. The test covers different domains, including visuospatial skills/executive function (5 points - trail making, cube copying, and clock drawing), naming (3 points - animal naming), attention (6 points - digit repetition, vigilance task, and serial subtraction), language (3 points - sentence repetition and phonemic verbal fluency), memory (5 points - delayed word recall), and temporal and spatial orientation (6 points), totaling a maximum score of 30 points [34]. The application will be performed individually, in a quiet and adequately lit environment, with the participant sitting comfortably in a chair with a backrest and armrests, positioned in front of a table, in order to ensure postural stability and standardization of the assessment conditions.

#### BESTest

Balance will be assessed through the Balance Evaluation Systems Test (BESTest), a comprehensive clinical instrument designed to identify specific balance system impairments underlying postural instability. The test evaluates six distinct balance domains: biomechanical constraints (5 items), stability limits/verticality (3 items), anticipatory postural adjustments (5 items), postural responses (5 items), sensory orientation (2 items), and stability in gait (7 items), totaling 27 items. Each item is scored on an ordinal scale ranging from 0 (severe impairment) to 3 (no impairment), with a maximum total score of 108 points, which can be expressed as a percentage of the total score. The assessment will be conducted individually in a quiet, well-lit environment, following standardized instructions. Participants will wear comfortable clothing and perform the tasks under the supervision of a trained examiner, ensuring safety and consistency throughout the evaluation [33].

#### Beck Depression Inventory (BDI)

Mood will be assessed using the Beck Depression Inventory (BDI), a widely used self-report instrument designed to evaluate the presence and severity of depressive symptoms. The BDI will be administered individually in a quiet and adequately lit environment, following standardized instructions. Total scores will be used as an indicator of depressive symptom burden and included as a covariate in adjusted analyses because mood may influence both cortical activation and behavioral performance during dual-task walking [35].

#### Assessment and Data Collection Workflow

After enrollment, participants will be stratified into three groups: young adults, healthy older adults, and individuals with PD. At baseline, all participants will undergo standardized clinical and functional assessments, including demographic characterization, cognitive screening with the MoCA, and balance evaluation with the BESTest. They will then complete the experimental protocol, consisting of motor single-task walking, cognitive single-task verbal fluency, and dual-task walking, while cortical hemodynamic activity and spatiotemporal gait parameters are simultaneously recorded using fNIRS and the GAITRite system.

The same assessment protocol will be repeated annually over 5 years, resulting in five assessment time points: baseline and four annual follow-ups. At each follow-up, participants will undergo the same sequence of clinical, cognitive, and experimental assessments, allowing longitudinal evaluation of changes in cortical activation, gait performance, and motor and cognitive dual-task costs over time.

An overview of participant stratification, assessments, and experimental procedures is shown in Figure 4.

**Figure 4.**
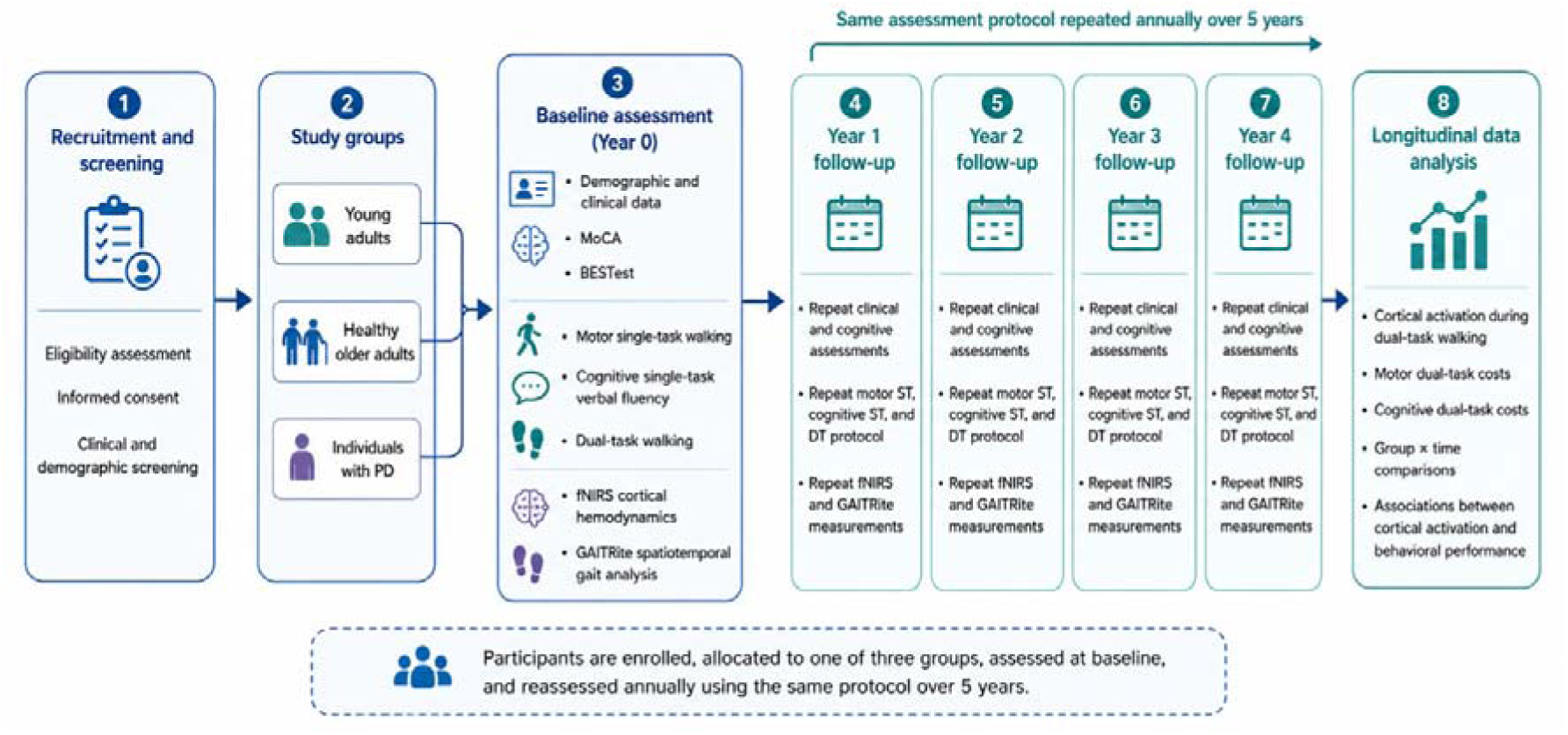
Study flowchart. Figure 4 illustrates the study workflow showing recruitment, group allocation, baseline assessment, four annual follow-ups, and longitudinal data analysis. Young adults, healthy older adults, and individuals with PD will undergo the same clinical, cognitive, and experimental protocol at baseline and at each yearly follow-up, including MoCA, BESTest, motor single-task walking, cognitive single-task verbal fluency, dual-task walking, and simultaneous fNIRS and GAITRite assessments. Longitudinal analyses will examine cortical activation during dual-task walking, motor and cognitive dual-task costs, and their associations across groups over time.

### DATA SOURCE

All data will be derived from primary data collection conducted specifically for this study. Clinical, cognitive, neurophysiological, and gait-related data will be acquired prospectively from participants during standardized laboratory-based assessments. Demographic and clinical information will be collected through structured interviews and standardized clinical instruments.

### STUDY SIZE

The sample size was determined considering the longitudinal design of the study and the primary outcome measure of interest, namely the change over time in cortical activation during dual-task walking across groups. The primary effect of interest will be the group × time interaction, testing whether trajectories of cortical activation differ across young adults, healthy older adults, and individuals with PD over 5 years.

Because longitudinal effect-size estimates for annual changes in cortical activation measured with fNIRS across these three groups remain limited, the study was planned to ensure an analyzable final sample of 31 participants per group at the last assessment. Assuming an annual attrition rate of 15% and a total of five assessments (baseline plus four annual follow-ups), the expected retention at the final assessment was estimated at 52.2%. Based on this retention rate, the required baseline sample was estimated at 60 participants per group, resulting in a total sample of 180 participants.

This sample size is expected to provide adequate support for the planned longitudinal mixed-effects analyses of the group × time interaction, while also accounting for losses related to withdrawal, clinical progression, and technical issues in repeated fNIRS assessments.

### QUANTITATIVE VARIABLES / STATISTICAL METHODS

The primary outcome measure will be the longitudinal change in cortical activation during dual-task walking, quantified by HbO, across prespecified prefrontal and premotor cortical regions.The main predictors of interest will be motor and cognitive dual-task costs derived from gait and verbal fluency performance across experimental conditions. Covariates will include age, sex, education, cognition, balance, mood, and, in the PD group, disease severity.

Baseline characteristics will be summarized using descriptive statistics. Continuous variables will be presented as mean and standard deviation or median and interquartile range, according to distribution, and categorical variables as absolute and relative frequencies.

Longitudinal analyses will be performed using linear mixed-effects models to account for repeated observations within participants over time. Cortical activation will be modeled as the primary dependent variable. The main parameters of interest will be group, time, motor dual-task cost, cognitive dual-task cost, and the relevant interaction terms, particularly those testing whether longitudinal changes in cortical activation and its association with behavioral performance differ across young adults, healthy older adults, and individuals with PD. Participant-specific random intercepts will be included to account for within-subject dependence, and random slopes for time will be considered when supported by model fit and convergence. Time will be modeled primarily as a categorical variable to allow non-linear trajectories across annual assessments; supplementary models treating time as continuous may be explored to estimate linear rates of change.

Adjusted models will include age, sex, and education a priori. Additional adjusted models may include cognition, balance, and mood. In the PD group, disease severity measures will be considered in exploratory adjusted analyses. Baseline gait performance may also be included, when appropriate, to improve interpretation of longitudinal associations between cortical activation and behavioral performance.

Additional mixed-effects models will be fitted to examine motor and cognitive dual-task costs as dependent variables, with cortical activation entered as a predictor, in order to explore the bidirectional relationship between neural recruitment and behavioral performance over time. These analyses will be considered complementary to the primary models.

Prespecified prefrontal and premotor cortical regions will be examined to account for the distributed cortical control of gait under dual-task demands. For analyses involving multiple cortical regions or multiple complementary outcomes, correction for multiple comparisons will be applied using the false discovery rate approach. Findings will be interpreted considering the multiregional nature of cortical recruitment during dual-task walking. Model assumptions will be evaluated by inspection of residuals, homoscedasticity, and influential observations. If necessary, robust estimation procedures or appropriate transformations will be considered.

Missing data will be handled within the mixed-model framework under the assumption that data are missing at random, allowing inclusion of participants with incomplete follow-up. The amount and pattern of missing data will be described for each annual assessment. Sensitivity analyses will include models restricted to participants with greater follow-up completeness and, when appropriate, alternative covariance structures or adjustment sets.

All tests will be two-tailed, with statistical significance set at 0.05. Effect estimates will be reported with 95% confidence intervals. Statistical analyses will be performed using appropriate software for longitudinal modeling.

## DISCUSSION

This longitudinal protocol provides a standardized framework to investigate how cortical activation during dual-task walking relates to concurrent motor and cognitive performance across young adults, healthy older adults, and individuals with PD. By combining annual repeated assessments with the same experimental paradigm, the study is designed to move beyond cross-sectional descriptions and examine whether age- and disease-related changes in gait automaticity are accompanied by distinct trajectories of cortical recruitment over time [3,5]. This is a central unresolved question in the field, as current evidence still does not clarify whether increased cortical activation during dual-task walking reflects adaptive compensation, reduced neural efficiency, or stage-dependent ceiling effects [3,5,16].

The protocol is particularly relevant because dual-task walking is an ecologically valid model of locomotor control under real-world demands and because dual-task costs are consistently greater in aging and PD than in single-task walking [4,5,15,36]. However, most previous studies have relied on cross-sectional designs, pairwise group comparisons, and predominantly prefrontal measures, limiting the interpretation of how cortical activation relates to concurrent behavioral performance [12,15,20]. In addition, longitudinal evidence remains scarce, especially for repeated cortical measures during dual-task walking in PD and for direct comparison of healthy aging and PD under the same experimental framework [21,23,24,25]. In this context, the present study has strong potential to generate novel evidence by examining how cortical activation and behavioral dual-task costs evolve over time in different populations exposed to the same task demands.

A major strength of the protocol lies in its design. Applying the same dual-task paradigm to young adults, healthy older adults, and individuals with PD improves comparability across groups and reduces the interpretive limitations of non-equivalent protocols [5,10,13]. The inclusion of separate motor and cognitive single-task conditions supports a more precise interpretation of dual-task-related changes by distinguishing neural activity related to gait, cognition, and their interaction [13,14]. Repeated blocks, randomized order, broader cortical coverage including prefrontal and premotor regions, and simultaneous assessment of multiple spatiotemporal gait parameters further strengthen the protocol and may improve the reliability and interpretability of both hemodynamic and behavioral measures [19,20,26]. Together, these features make the study well suited to examine whether longitudinal changes in cortical recruitment are accompanied by worsening, preservation, or reorganization of motor and cognitive performance under dual-task demands.

The longitudinal component is one of the most original aspects of this study. Existing longitudinal work has mainly focused on behavioral dual-task gait outcomes in older adults, including prediction of falls and cognitive decline, or on longitudinal changes in prefrontal activation during dual-task walking in healthy older adults [21,23,24,25]. By contrast, repeated cortical assessment during dual-task walking in PD remains limited, and direct comparison of age-related and PD-related trajectories is still scarce [24,25]. This protocol therefore offers an opportunity to clarify whether cortical recruitment follows different trajectories in healthy aging and PD, and whether these trajectories are associated with distinct patterns of motor and cognitive dual-task cost. Such evidence may help refine current models of reduced gait automaticity and provide a more mechanistic interpretation of cortical activation during dual-task walking.

The findings generated by this study may also have important translational implications. A better understanding of how cortical activation patterns relate to dual-task motor and cognitive performance may help identify clinically meaningful neural signatures of declining gait automaticity. This may contribute to the development of more targeted rehabilitation strategies aimed at improving cognitive–motor integration during walking and at tailoring training according to disease stage and functional profile. In addition, more precise characterization of the cortical regions and activation patterns associated with more favorable or less favorable dual-task performance may help guide future therapeutic approaches involving non-invasive brain stimulation, particularly those targeting frontal and premotor regions to modulate gait-related cognitive-motor control. Although the present study is not interventional, it may provide mechanistic evidence that supports the selection of candidate targets and outcomes for such interventions.

Some limitations should be acknowledged. fNIRS is restricted to superficial cortical regions and cannot directly assess subcortical locomotor networks, which are highly relevant to gait automaticity in aging and PD [19,31]. As in other dual-task paradigms involving speech, phonemic verbal fluency may introduce physiological and movement-related noise, although the protocol includes short-separation channels, repeated blocks, and standardized preprocessing procedures to reduce these effects [20,26]. Attrition and variability in follow-up completeness are also inherent challenges in longitudinal designs. Nevertheless, by integrating repeated neural, motor, and cognitive assessments within a standardized framework, this protocol is well positioned to advance current understanding of how cortical activity during dual-task walking changes across healthy aging and PD.

Taken together, this study has the potential to generate new evidence on the role of cortical activation during dual-task walking by examining how cortical and behavioral dual-task costs change over time in healthy aging and PD. By clarifying how modifications in cortical activity patterns relate to motor and cognitive performance, the protocol may strengthen the mechanistic basis for future rehabilitation and neuromodulation strategies grounded in cognitive-motor control.

### KEY RESULTS and INTERPRETATION

Not applicable.

### BIAS and LIMITATIONS

Potential sources of bias and limitations should be considered when interpreting the findings of this protocol. fNIRS captures activity only from superficial cortical regions and therefore cannot directly assess subcortical networks involved in gait automaticity [3,19,32]. Although the protocol includes short-separation channels, repeated blocks, and standardized preprocessing procedures, residual motion-related and physiological artifacts may still affect cortical signals during walking [20,26]. Participants will be recruited through convenience sampling, which may limit representativeness and introduce selection bias. Additional variability may arise from individual differences in task prioritization, motivation, fatigue, and verbal output strategies, as well as from clinical heterogeneity within the PD group. As a longitudinal study, the protocol is also subject to attrition and incomplete follow-up over time, which may reduce precision in trajectory estimates despite the use of mixed-effects models.

### GENERALIZABILITY

The findings generated by this protocol may have important clinical and translational implications for the assessment of cognitive-motor interference during gait across healthy aging and PD. By integrating spatiotemporal gait parameters with concurrent measures of cortical activation in prefrontal and premotor regions during overground walking, this study may help identify neural and behavioral markers of reduced gait automaticity that are not evident under single-task conditions. Such knowledge may inform more targeted rehabilitation strategies and support future therapeutic approaches involving non-invasive brain stimulation aimed at modulating gait-related cognitive-motor control.

## CONCLUSION

In summary, this protocol provides a standardized longitudinal framework for investigating cognitive-motor interactions during walking through the integration of behavioral gait measures and concurrent cortical activity. By examining how cortical and behavioral dual-task costs evolve over time across healthy aging and PD, the study may advance understanding of reduced gait automaticity and support the future development of more precise rehabilitation and neuromodulation strategies.

## DECLARATIONS

### Ethics approval and consent to participate

The study was approved by the Research Ethics Committee of the Hospital das Clínicas, Faculty of Medicine, University of São Paulo (CAAE: 67388816.2.0000.0065; Approval number 6.913.344). All participants will be informed about the study procedures and objectives prior to participation and provided written informed consent before the assessments will be conducted.

### Consent To Publish

Prior to the beginning of the assessments all participants will be informed about the research and sign an informed consent form, authorizing the analysis and publication of the data obtained from their participation.

### Data Availability

The datasets generated and/or analyzed during the current study are not publicly available but will be available from the corresponding author on reasonable request.

### Author Contributions

MEPP and JRS designed the study. MEPP, JRS, PRS, LBRS and LMA designed the assessment protocol. Statistical analysis will be performed by MEPP, JRS and KRB. The draft of the manuscript was prepared by LMA, PRS, GVS and DFG and was critically reviewed by MEPP and JRS.

## ACKNOWLEDGMENTS

This paper is dedicated to the memory of Gabriel Venas Santos, whose unwavering commitment to this project and to our research group will continue to inspire this work for years to come.

The authors sincerely thank all participants for their collaboration and commitment, as well as the technical and administrative teams who provided essential support in carrying out the research activities. We also acknowledge the institutions involved for their continuous support in the development of this work.

